## Supplementary Tables and Figures for "Alcohol consumption is associated with structural changes in various organ systems: A population-based study in UK Biobank"

**Supplementary Table 1. Alcohol consumption (g/d) in all and males and females for brain grey matter (N=10143), brain white matter (N=9053), heart (N=11821) aorta (N=12376) and liver (N=3649).**

| **Alcohol (g/d)** | **Brain grey matter** | **Brain white matter** | **Heart** | **Aorta** | **Liver** |
| --- | --- | --- | --- | --- | --- |
| **All (median)** | 14.29 | 14.29 | 14.29 | 14.29 | 16.61 |
| **IQR** | 6.46-26.78 | 6.26-26.79 | 6.70-26.79 | 6.69-26.79 | 8.93-28.86 |
| **Males (median)** | 20.94 | 20.97 | 21.43 | 20.99 | 23.52 |
| **IQR** | 10.28-35.78 | 10.27-35.75 | 10.71-36.60 | 10.28-36.19 | 12.93-38.79 |
| **Females (median)** | 10.71 | 10.71 | 10.71 | 10.71 | 12.47 |
| **IQR** | 3.57-17.89 | 3.57-17.86 | 3.99-18.58 | 3.89-17.31 | 6.80-21.43 |

IQR: Interquartile range

**Supplementary Table 2. Coefficients for log_2_ alcohol and age in an expanded model for cardiac (N=11,821) and aortic (N=12,376) imaging phenotypes including tests of interactions between log_2_ alcohol and age.**

|  | **Alcohol** | **Alcohol x Age** | **Age** |
| --- | --- | --- | --- |
| **Heart** | ***Beta, P-value*** | ***Beta, P-value*** | ***Beta, P-value*** |
| Left ventricular mass index | 1.84, 3.2×10^-10^ | -0.02, 3.5×10^-7^ | -0.01, 0.42 |
| Left ventricular end-diastolic volume index | 2.25, 6.9×10^-5^ | -0.03, 3.5×10^-3^ | -0.24, 3.4×10^-11^ |
| Left ventricular ejection fraction (%) | 0.17, 0.50 | -0.002, 0.62 | 0.04, 6.8×10^-3^ |
| Right ventricular end-diastolic volume index (ml/m^2^) | 1.99, 1.0×10^-3^ | -0.02,0.018 | -0.25, 4.7×10^-11^ |
| Right ventricular ejection fraction (%) | -0.04, 0.87 | 0.001, 0.72 | 0.04, 0.024 |
| Left atrial volume index (ml/m^2^) | 0.59, 0.23 | -0.002, 0.75 | -0.11, 4.4×10^-4^ |
| Right atrial volume index (ml/m^2^) | 0.25, 0.65 | 1.4x10^-4^, 0.99 | 0.04, 0.22 |
| **Aorta** |  |  |  |
| Ascending aortic area index (mm^2^/m^2^) | 7.4, 0.06 | -0.08, 0.22 | 3.0, 1.5×10^-34^ |
| Descending aortic area index (mm^2^/m^2^) | 5.9, 6.9×10^-4^ | -0.07, 8.1×10^-3^ | 2.1, 2.9×10^-83^ |
| Ascending aortic distensibility (%/mmHg) | -0.21, 1.4×10^-7^ | 0.003, 2.6×10^-7^ | -0.1, <1.0×10^-300^ |
| Descending aortic distensibility (%/mmHg) | -0.20, 4.3×10^-6^ | 0.003, 2.7×10^-5^ | -0.10, 6.0×10^-293^ |

Regression model: Imaging phenotype=log_2_ Alchohol + log_2_Alcohol* Age + Age + Sex + Ethnicity + BMI + Hypertension + Diabetes + Ever smoked + College degree

**Supplementary Figure 1**. **Flow chart of eligible participants included in analyses.**

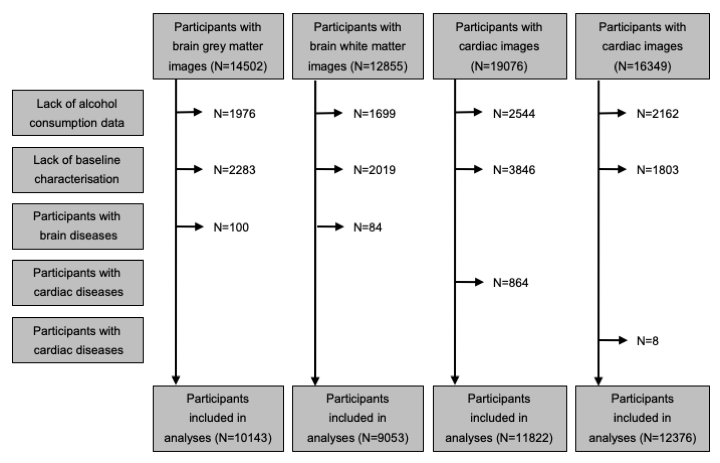

**Supplementary Figure2. Partial residual plots for the imaging derived phenotypes**

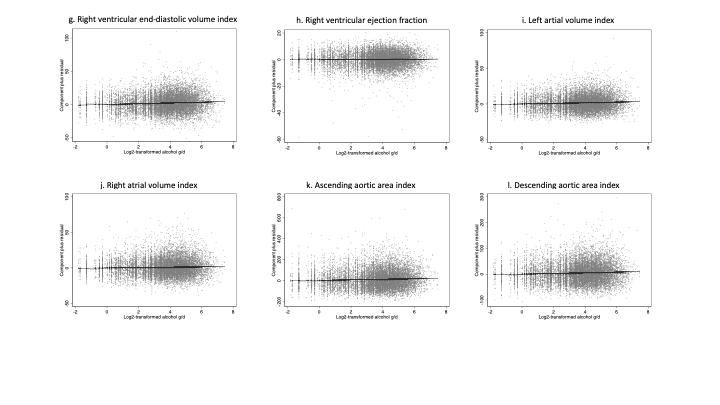

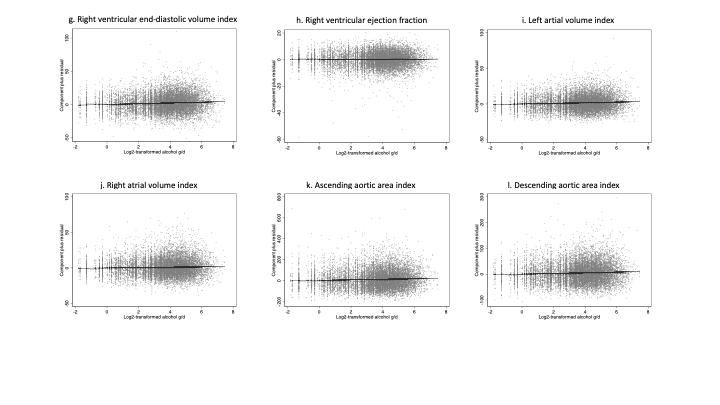

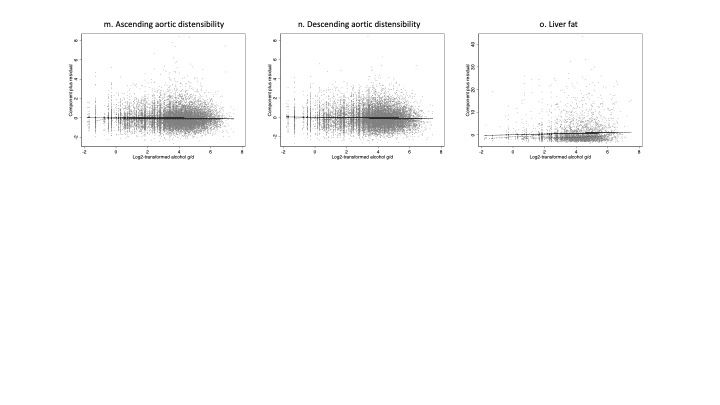
